# The genetics of idiopathic intracranial hypertension (IIH): Integration of population studies and clinical data

**DOI:** 10.1101/2023.06.03.23290934

**Authors:** Roei Zucker, Michael Kovalerchik, Zvika Davidovich, Ehud Banne, Idit Maharshak, Michal Linial

## Abstract

**Background:** Idiopathic intracranial hypertension (IIH) is a condition characterized by increased intracranial pressure without a known cause. IIH mainly affects overweight and reproductive-age women. Due to elevated intracranial pressure in IIH patients, papilledema (PAP), a disease caused by swelling of the optic disc, often co-occurs. Previous genome-wide association studies (GWAS) of common variants failed to find significant associations.

**Methods:** We applied genetic association protocols to a cohort of 173 patients diagnosed with IIH or PAP from the UK Biobank (UKB). The results were compared to the FinnGen data. We applied routine and coding GWAS (cGWAS) to a unified cohort of IIH/PAP. We also employed SKAT, which considers all variants within a given segment, and PWAS, which estimates the damage of variants to protein function and models a gene by aggregating its coding variants.

**Results:** The detection power of standard GWAS methods is restricted by cohort size and false discovery. To overcome these limitations, we considered gene-centric approaches for the unified group of IIH and PAP patients. Notably, the dominant comorbidity of IIH is PAP in the UKB and FinnGen populations. Seeking shared genes by GWAS for IIH and PAP identified VLDLR and SHANK2 genes in the UKB and FinnGen cohorts, respectively. By utilizing complementary gene-centric association protocols (coding GWAS, SKAT, and PWAS) for the unified IIH/PAP group, we identified 16 genes that were recognized by at least two methods, with FOXF1 and RGCC recognized by all three. Employing a functional enrichment scheme emphasized the significance of cilium, microtubule, and cytoskeletal functions. To gain insight into IIH etiology, we focused on the choroid plexus, a brain structure that produces and secretes the cerebrospinal fluid (CSF). Altogether, 7 of the 16 candidate genes are listed among the 281 genes with enhanced expression in the choroid plexus epithelium. Among them, MAPK15, DNAH5, and SLC28A3 are involved in ciliary microtubule dysregulation.

**Conclusions:** This study highlights the strength of integrative genetic association approaches with functional and clinical knowledge. By identifying potential genetic effects, we propose the biological and cellular relevance of the choroid plexus for IIH etiology and suggest candidate genes for further investigation.

## Background

Idiopathic intracranial hypertension (IIH, also called p*seudotumor cerebri*) is a condition of raised intracranial pressure (ICP) with no proven pathogenesis. It is linked to obesity, and patients are typically overweight women of reproductive age (1). Three chief intracranial mechanisms have been proposed to cause raised ICP, including disorderly cerebrospinal fluid (CSF) dynamics such as CSF hypersecretion, outflow obstruction, and increased venous sinus pressure. Whereas the pathogenesis of IIH is unknown, dysregulation of ICP is an important direction of investigation.

From an epidemiological perspective of IIH, the annual incidence is 1-2 per 100,000 individuals, with a much higher incidence for females at their reproduction age (2). The prevalence of obese women at the age of 20 to 44 years is approximately ∼20 cases per 100,000 (3). Patients with IIH are usually diagnosed based on the updated modified Dandy criteria (4), which necessitate the existence of a raised ICP, papilledema, and no identifiable secondary causes. In a study that compared cases with IIH and papilledema, only 5.7% of patients diagnosed with IIH did not have papilledema (5). In the past, there was little evidence to guide the management of IIH, but significant prospective studies have contributed to our understanding of the disorder. Examples of such studies include the Idiopathic Intracranial Hypertension Treatment Trial (IIHTT), which examined the effects of the carbonic anhydrase inhibitor drug acetazolamide in patients with mild visual loss (6). Additional studies tested the effect of a low-calorie diet on ICP and symptoms and signs of IIH (7). A revised nomenclature for IIH was proposed for a more accurate patient classification (8). Still, the pathogenic mechanisms of IIH are uncertain.

BMI, sex, age, ethnicity, and environmental factors (e.g., nutrition) are known risk factors for IIH. Preliminary findings proposed an improvement in symptoms and signs of IIH with weight reduction (9). This improvement was not established until investigators of a prospective cohort study (7) tested for significant improvements in ICP, papilledema, and headache after weight loss compared with a control period before weight loss. The mechanism by which weight loss improves IIH is uncertain. The effect of weight in terms of mass alone is a weak explanation because BMI and lumbar puncture opening pressure have a frail, non-significant relationship (10). Glucagon-like peptide 1 (GLP-1) receptor agonists emerge as new metabolic therapeutic targets for IIH (11). Recall that GLP-1 receptor agonists (RAs) are extensively used in the treatment of type 2 diabetes mellitus, and, recently, also in managing obesity (12). In the clinical view of IIH, the usage of GLP-1 RAs ranges beyond weight reduction. Importantly, GLP-1 receptors have been recognized to be expressed in the choroid plexus epithelium of humans and rodents. When stimulated with exenatide, a GLP-1 RA drug, it reduces CSF secretion. It has been shown that rats with intracranial hypertension that were treated with exenatide exerted a lowered morbidity with a lowering of the high ICP by 50%, with a persistent effect that lasted over a week (13).

Similar to other complex diseases, we anticipate that the genetic landscape of individuals also affects their susceptibility to IIH. Such genetic effects are likely to be associated with IIH comorbidity of vision complications, specifically papilledema (PAP). Due to the complex nature of the syndrome and its rarity, no causal variants were identified. A recent genome-wide association study (GWAS) analysis was performed on female patients from the IIHTT cohort and their matched controls (95 women in each group). Altogether, from approximately 300K high-quality common SNPs, 15 associations were reported with weak statistics (p-values ranging from 1e-07 to 1e-05) (14). Revisiting the results from FinnGen Fz8 for the group of patients with IIH (n = 197) or papilledema (PAP; n = 464) also failed to report associated variants that met the statistical threshold.

The rate of false positive discoveries underscores the identification of the genetic basis of complex diseases with a limited number of patients and a lack of strong heritability. Therefore, biological insight and mechanistic interpretation are often limited. In addition, about 90% of the associated variants are not assigned to any obvious causal gene (15). In this study, we sought IIH/PAP genetics in the UK Biobank (UKB) by applying complementary state-of-the-art gene-based approaches. The goal of this study is to explore genetic signals for IIH/PAP, and identify the top candidate genes that are significantly associated with these conditions. We also analyzed the comorbidity of IIH/PAP and discussed the impact of current genetics and medical records on patients’ care.

## Methods

### Data resource and processing

The IIH is indexed by OMIM #243200. The other indices are ICD-9 (International classification of diseases, ninth revision) term 348.2, the OrphaNet #238624 and the disease ontology (DO) 11459. We used the UK Biobank (UKB) database for identifying the individuals with the selected ICD-10-CM classification, using main or secondary diagnoses fields (UKB data-fields 41202 and 41204, respectively). We restricted the analysis to ICD-10 combined phenotype of G93.2 (Benign intracranial hypertension) and H47.1 (Papilledema). We used the genetic ethnic grouping, (data-field 22006) to remove genetic relatives, by randomly keeping only one representative of each kinship group.

For the GWAS results from FinnGen Fz9, we extracted the reported analyses for the endpoint outcomes of IIH (Benign intracranial hypertension, G6_BENINTRAHYP), and PAP (Papilloedema, unspecified; H7_PAPILLOEDEMA). FinnGen Fz9 consists of >377,000 individuals, >20 M variants, and 2,272 characterized diseases. Note that the FinnGen relies on high imputation accuracy down to very low allele frequencies (AFs), which enabled the identification of associations with low-frequency variants using a routine GWAS approach (16).

### Genetic Data

We tested 804,069 informative markers that were represented in UKB cohort. We considered only variants that are included in the 18,053 coding genes (including splicing variants). This set included the 639,323 coding genes imputed variants that were selected from the 97,013,422 whole genomes UKB imputed set. To select current knowledge from GWAS resources, we have used the Open targets genetic platform, (OTG, dated 3/2022) (17). The OTG compiled information on any GWAS performed for IIH (EFO_1001132) and its synonymus phenotypes (e.g., benign intracranial hypertension).

### Association tests

We performed four different association tests coined all GWAS, coding GWAS, SKAT and PWAS. For all methods, we included 8 covariates including sex, year of birth, and the first 6 principal components (PC) to account for population structure.

#### All GWAS

It refers to the routine GWAS protocol with ∼97M imputed variants (according to UKB imputation protocol (18)). We applied the standard PLINK protocol to filter candidate variants for the GWAS analysis. We considered variants with a MAF threshold of 0.001, Hardy-Weinberg equilibrium (HWE) exact test of p-value 1e-6, and genotyping coverage with 90% call rate. Altogether, we analyzed 10,258,628 variants. We also included as covariates sex, year of birth and the first six principal components (PCs) to account for population structure. We only considered variants within genes (exons and introns) as indicator for gene association and have not applied variant to other functional units beyond coding genes. The IIH/PAP group includes 173 patients and 291,139 controls. For IIH and PAP we included 121 and 100 patients, with 291,207 and 291,228 as controls, respectively. There are 16 patients with both diagnoses. These patients were not included for the specific GWAS, but were included for analyzing the IIH/PAP group.

#### Coding GWAS

It refers to a restricted GWAS, where the analysis covers variants in the coding regions of genes. Altogether, the coding region set contained 639,323 variants, covers ∼1% of the genome. To improve reliability, we filtered out variants by minor allele counts (MAC <20). Coding GWAS allows greater interpretability of the gene results since the connection of variant and function is plainer (19).

#### SKAT (sequence kernel association test)

SKAT is a collapse-based association method that allows the estimation of the correlation of multiple variants at the same time to a phenotype. This approach allows to account for the combined effect of variants according to the preselection of the relevant genomic information (e.g., gene). SKAT was applied with the same covariates as listed above. We provided SKAT with the genotypes of all protein-imputed variants by the additive genotypes. We have used version 2.2.5 of the R package of SKAT (20), with output type for a dichotomous phenotype. For the gene-based SKAT, we used an adjusted p-value following Bonferroni correction.

#### PWAS (proteome-wide association study)

PWAS is a genetic analysis method that estimates the effect of variations on the protein-level. Using the machine learning model of FIRM (21), the effect of each person’s variants on protein function is estimated. Like SKAT, PWAS uses an aggregation method to analyze the genetic effect on phenotypes. PWAS aggregates the effect scores into a single score for each gene. To find the effect and significance of each gene’s score on the phenotype, we used logistic regression. We ignore models that were designed to address dominant or recessive heritability (21). The corrected p-value is by applying FDR <0.05.

### Functional enrichment

STRING platform was used for functional enrichment by applying a high connectivity network threshold (at a protein-protein confidence score of 0.7). For gene expression analysis in the developing brain, we used FUMA-GWAS application of Gene2Func (22). It was also used by default parameters for functional enrichments from a list of genes, with all coding genes as the background gene set (22). We have used the Enrichr KB platform (23) to identify the genes that are significantly connected to GWAS terms and HPO terms. We only considered the top 10 results for each GWAS report as candidate genes for the knowledge-based representation. Enrichr uses hypergeometric consideration for p-value and its adjusted value by correcting for multiple hypotheses.

A collection of ‘tissue-enhanced’ gene expression of choroid plexus extracted from HPA (human proteome atlas) compilation (24). About 65% of the human proteome is expressed at some level in the choroid plexus. However, we considered the differentially expressed subset of 281 enhanced genes (24).

## Results

### IIH from the UK biobank (UKB)

Large population cohorts are routinely used for studying common complex diseases and human traits. In this study, we address the rare phenotype of IIH by genetic association and analyzing IIH comorbidities. We limited the analyses to European origin without family members with (see Methods). A substantial number of patients with IIH may also suffer from unspecified papilledema (PAP), we inspected both related conditions.

**Table 1** summarizes the UK biobank (UKB) demographic statistic of IIH and PAP patients. We considered individuals of European origin, after excluding genetic relatives, and poor genotyping data (see Methods).

**Table 1.** IIH/PAP cohort statistical properties.

| Measure | Group | Males | Females | Both |
| --- | --- | --- | --- | --- |
| Number | IIH/PAP | 70 | 103 | 173 |
|  | IIH | 38 | 67 | 105 |
|  | PAP | 35 | 49 | 84 |
|  | (IIH+PAP) | 3 | 13 | 16 |
| Age of diagnosis | IIH | 42.50 | 39.60 | 40.00 |
|  | PAP | 46.00 | 36.40 | 38.20 |
| BMI (mean) | IIH/PAP | 29.03 | 30.91 | 30.14 |
|  | IIH | 29.21 | 32.25 | 31.15 |
|  | PAP | 29.45 | 29.77 | 29.64 |
|  | (IIH+PAP) | 36.22 | 33.61 | 34.10 |
| BMI (s.d.) | IIH/PAP | 4.94 | 6.70 | 6.10 |
|  | IIH | 5.47 | 7.06 | 6.66 |
|  | PAP | 5.60 | 5.55 | 5.54 |
|  | (IIH+PAP) | 12.97 | 4.91 | 6.54 |

We have created a unified cohort of individuals that were diagnosed with idiopathic intracranial hypertension (IIH, ICD-10: G93.2) or with unspecified papilledema (PAP, ICD-10: H47.1). There are 173 IIH/PAP individuals. We observed a very significant overlap of these two rare diseases (IIH+PAP, n=16, hypergeometric test p-value = 1.471e-39). There are 60% of females in the IIH/PAP group and a higher fraction of females (81%) among the subgroup of the overlapping diagnoses. Statistical tests for the fraction of females within the IIH group relative to the entire female population confirmed their enrichment (Mann-Whitney U-test, p-value: 0.0046). Similarly, females are enriched when IIH and PAP groups were compared (U-test, p-value: 0.0252). Interestingly, none of these tests were significant for males (Supplementary **Table S1**).

We also observed that females are diagnosed at an earlier age relative to males. The age of diagnosis in males is about 3 and 10 years later for IIH and PAP, respectively (**Table 1**). Inspecting the subset of the 16 individuals with both diagnoses (denoted as IIH+PAP) indicated that for the majority (11/16) the diagnosis was provided at the same time, while for 4 of the other 5 cases, IIH diagnoses preceded PAP. High BMI characterized both females and males, with a significant difference in the sex distributions (Supplementary **Fig. S1**). Independent cohort from FinnGen resource corroborated these observations. However, the ratio of females to males with IIH is much higher (86%, total 358 patients) with earlier diagnosis age (age 32.9 relative to 39.8 for females and males, respectively).

### Comorbidities with IIH

In seeking links to the clinical implication of IIH and PAP patients, we tested the occurrences of other clinical ICD-10 diagnoses as comorbidities. To this end, we collected ICD-10 associated with each of the patient and assign statistics according to the expected overlap.

**Table 2** shows the ranked list of diseases in the UKB for patients with IIH and PAP groups. We report the high occurrence comorbidities (n>30). The most significant linked diseases are obesity, headache, and essential hypertension. We also observed an enrichment of usage of anticoagulants, circulation problems, type 2 diabetes, and depression. Comparing the finding to the independent cohort from the FinnGen by ICD-10 outcomes confined that the most significant overlapping diagnoses include obesity, migraine, hypertension, pain, and mental disorders. Notably, in the FinnGen data, the overlap of patients that are diagnozed with IIH and PAP is much higher (48%), supporting the clinical rational for IIH/PAP unification. Lists of the ICD-10 identified for the IIH/PAP cohort from UKB and FinnGen are available in Supplementary **Table S2**.

**Table 2.** IIH /PAP patients from UKB and comorbidities.

| ICD-10 | Disease name | p-value | Counts (n) |
| --- | --- | --- | --- |
| **G93.2** | **Benign intracranial hypertension** | 0.0E+00 | 105 |
| **H47.1** | **Papilledema, unspecified** | 1.8E-282 | 84 |
| R51 | Headache | 1.5E-30 | 48 |
| Z86.7 | Personal history of diseases of the circulatory system | 7.0E-10 | 43 |
| I10 | Essential (primary) hypertension | 7.9E-10 | 98 |
| F32.9 | Depressive episode, unspecified | 9.2E-09 | 34 |
| E66.9 | Obesity, unspecified | 1.7E-06 | 32 |
| Z92.1 | Personal history of long-term (current) use of anticoagulants | 1.3E-05 | 31 |
| E11.9 | Type 2 diabetes mellitus, without complications | 1.8E-05 | 32 |
| K44.9 | Diaphragmatic hernia without obstruction or gangrene | 3.5E-04 | 37 |
| E78.0 | Pure hypercholesterolaemia | 1.1E-03 | 43 |
| Z86.4 | Personal history of psychoactive substance abuse | 7.0E-03 | 42 |

### Association studies from large populations

**Fig. 1** presents the framework of the analyses that were performed in seeking associations from large population sources. We focus on participants from the UKB for applying routine GWAS (coined all GWAS), coding GWAS and gene-based PWAS and SKAT. We also report on summary statistics from FinnGen Fz9, a representative data source for a well-phenotyped isolated population (25). We revisit the results provided by FinnGen from GWAS for the endpoint outcomes of IIH and PAP. The number of associations according to the p-value threshold used, for each method, is reported.

**Fig. 1.**
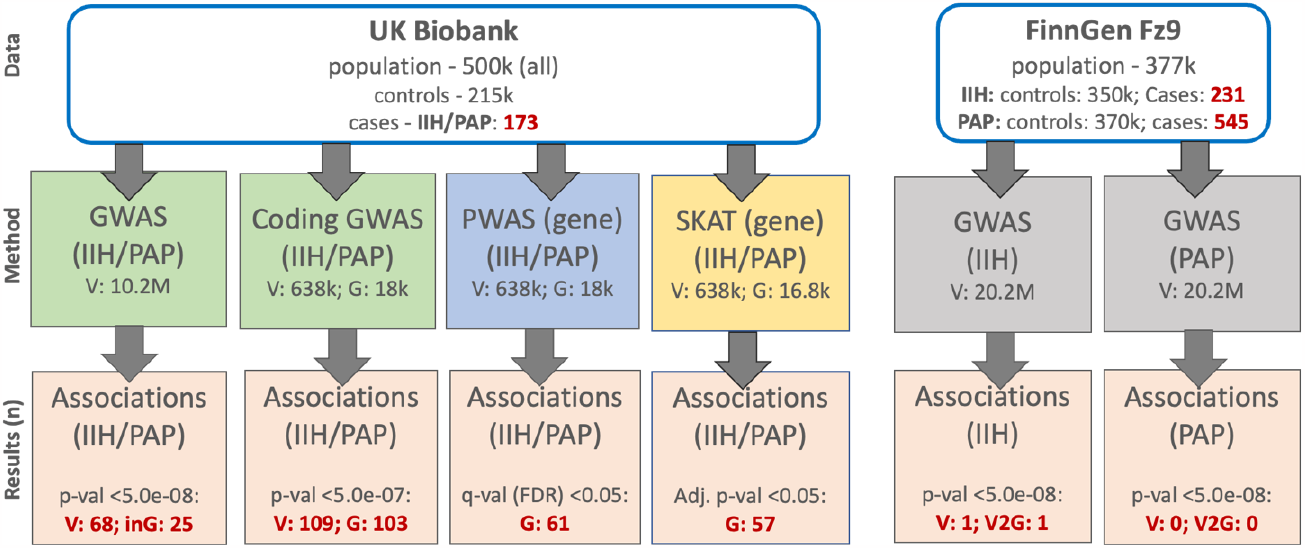
A scheme for the data source and methods for identifying associations from the UK Biobank (UKB) and FinnGen Fz9. The associated variants and related genes are reported for each method (in red). Note that for all GWAS the p-value threshold for significance is <5.0e-08. We report the number of variants within genes (i.e., variants within introns, 5’-UTR, and 3’UTR; inG). For coding GWAS, the accepted statistical threshold of full exomes was applied (p-value <5.0e-07). For gene-based methods, the resulting p-value is adjusted by Bonferroni correction or by FDR in the case of PWAS according to the number of genes analyzed. V, variant; G, coding gene; inG, within gene length.

### GWAS for the IIH/PAP group

We compiled a set of ∼97M variants to seek statistically relevant associations for the IIH/PAP group (173 patients). Following filtration (see Methods), we considered 10,258,628 variants which were partitioned to those located within gene length and intergenic ones. **Fig. 2A** shows a Manhattan plot for the IIH/PAP group. Among the 68 variants at p-value <5.0e-08 (green horizontal line, **Fig. 2A**), 28 variants are located within gene length (25 unique genes), where the rest are intergenic.

**Fig. 2.**
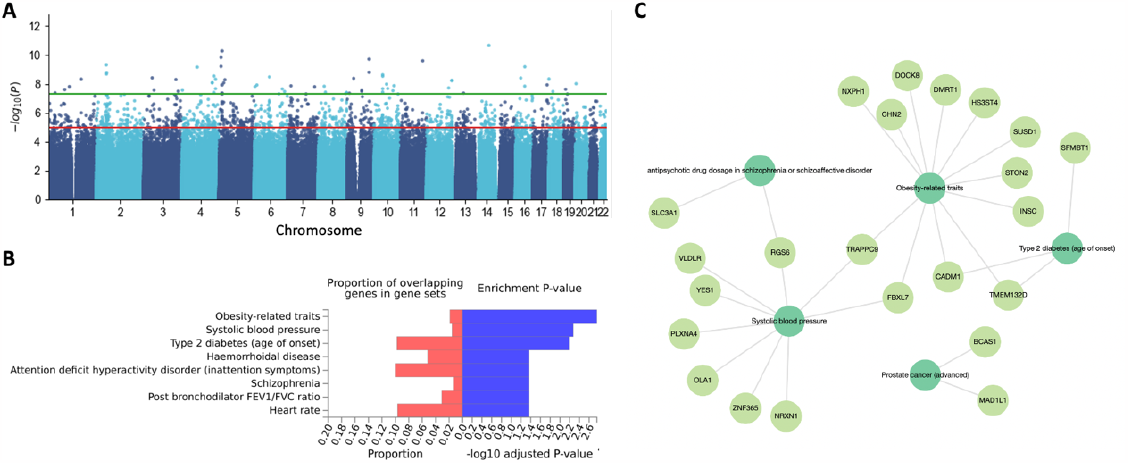
GWAS results of ∼10.2M variants for the IIH/PAP group. **(A)** Manhattan plot for Chr. 1 to Chr. 22. For visualization clarity we removed a single variant with extreme p-value (rs12203577, p-value 4.7E-70; Supplementary **Table S3**). The significant threshold of 5e-08 is depicted by the green horizontal line. **(B)** GWAS annotation enrichment of variants within the gene length based on the list of variants with p-value <5e-07 (69 genes). Enrichment visualization for main GWAS results is based on GWAS-FUMA Gene2Func protocol. **(C)** Gene-phenotype network extracted from the results of GWAS catalog according to functional enrichment of EnriChR KB (see Methods).

**Fig. S2**. shows Venn diagrams for all GWAS results at thresholds of p-value <5e-08 and <5e-08 with patients that are only diagnosed with IIH, PAP and the unified group (both; IIH/PAP including IIH+PAP, see **Table 1**). Only VLDLR (very low-density lipoprotein receptor) was shared by PAP and IIH with four supporting SNPs (9:2648868_A_T, rs74960704, rs543108298, rs577402784 and rs74960704; Supplementary **Table S3**). Interestingly, all 4 variants are rare or ultra-rare according to large population data. The supporting variants rs74960704, rs543108298, rs577402784 and rs956197858 are associated with MAF of 4e-03, 7e-06, 7e-06 and 8e-06. At a more relaxed threshold, also NXPH1 gene is shared between PAP and IIH groups with different variants governing the associations (Supplemental **Fig. S2**). Specifically, a single variant was reported for the IIH group, and 61 variants of NXPH1 were reported in the PAP group. The results of the overlapping groups are compiled in supplemental **Table S3**.

Functional enrichment of the genes that met the a lower p-value threshold (5.0e-07) shows an enrichment significance with respect to traits from the GWAS catalog for obesity, type 2 diabetes and more (**Fig. 2B**). **Fig. 2C** shows a network connecting significant associated genes according to the results from the GWAS catalog using EnrichR KB platform. The most connected phenotypes are related to systolic blood pressure, obesity and type 2 diabetes. These phenotypes appear as comorbidities for IIH (Supplementary **Table S2**).

We conclude that the results of all GWAS while focusing only on gene length are informative **(Fig. 2C)**. Note that about 36% of all tested variants occur within a gene length (exons and introns), with the majority of them occurring in introns. We therefore decided to explicitly analyze the coding regions, while considering the impact of rare variants.

### Coding GWAS for IIH/PAP group

Coding GWAS for the IIH and PAP at p-value <5e-07 resulted with 162 genes and 173 genes, respectively, However, at this threshold the number of associated variants for the IIH/PAP is only 109 (103 genes). Among the 103 associated genes, approximately 60% and 30% were shared by the IIH and PAP groups, respectively. Only 15 additional genes were reported from the unified group (Supplementary **Fig. S3**). There are 16 shared genes (Supplementary **Fig. S3**). One of the shared gene identified by all groups is PCDHGA1 which is a potential calcium-dependent cell-adhesion protein that may be involved in the establishment and maintenance of neuronal connections in the brain. We anticipated that the unified phenotype of IIH/PAP exposes overlooked genetic signal. The rest of the analysis considers only the results associated with the IIH/PAP group.

**Fig. 3A** shows the effect size (OR) along with the p-values for the more significant associated 143 coding variants (p-values <1.0e-06, Supplementary **Table S4**). We observed that only a small set of variants displayed significant statistics and an elevated risk for IIH/PAP (**Fig. 3A**, shaded area). STRING analysis for functional enrichment for the associated genes showed a borderline protein-protein interaction (PPI) network significance (p-value 0.01). While no functional terms were significantly enriched, the network was manually partitioned into sub-clusters (with at least 3 interacting genes in each) that include vesicle trafficking, motor activity, calcium regulation, lipoproteins and microtubule-related activity (**Fig. 3B**). We anticipate that the underlying biology for IIH/PAP is complex and is likely not associated with a dominant pathway or cellular process

**Fig. 3.**
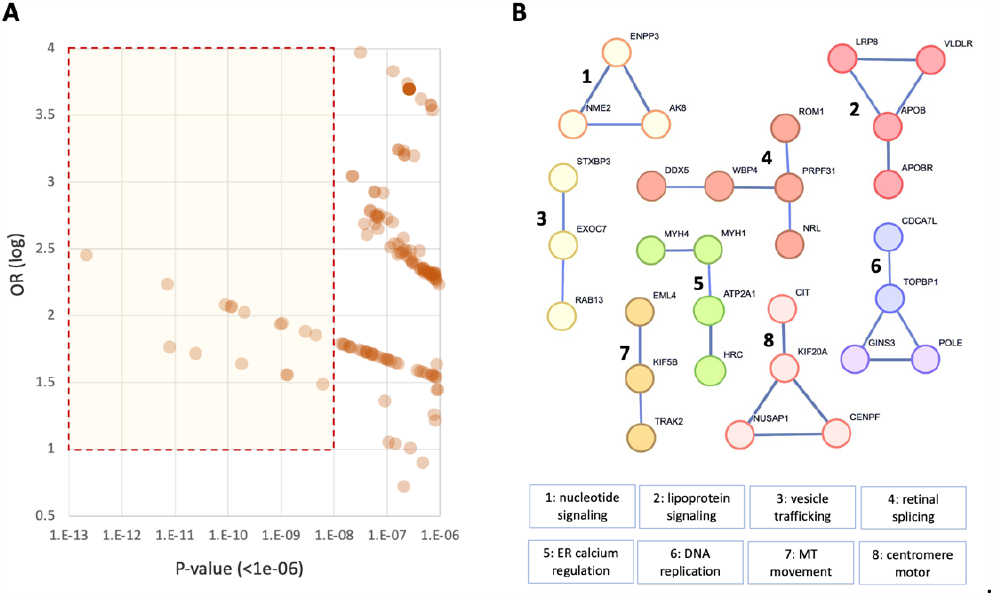
Coding GWAS associations to IIH/PAP. **(A)** Scatter plot of genes with p-value <1.0e-06 along with the measured odds ratio (OR). Genes with maximal effect size and statistical significance are included in the shaded area. **(B)** A network graph based on STRING results for the significant genes (<5e-06) according to coding GWAS for IIH/PAP group in the UKB. The STRING protein-protein interaction (PPI) confidence score of 0.7 was used. The network (with at least 3 connected genes) is partitioned into 8 clusters. Each numbered subgraph is indicated with its main functional property. MT, microtubule.

The most significant variants (p-value <5.0e-9) and their gene information are listed in **Table 3**. We report on the minor allele frequency (AF in %), and the consequence on the associated protein. In all cases, AF is below 0.0001 for the alternative (Alt) variant. Recall that the impact on protein function for rare variants can we substantial. For example, the gene PCDH15 is associated with a loss of function mutation (NP_149045.3: C1663fs).

**Table 3.** Properties of the 15 most significant coding GWAS variants (p-value <5e-09)

| ID | Ref | Alt | AF (%) | P-value | Gene | Name | Protein: consequence | Group<sup>a</sup> |
| --- | --- | --- | --- | --- | --- | --- | --- | --- |
| rs775238614 | CA | C | 0.0300 | 2.13E-13 | PCDH15 | protocadherin related 15 | NP_149045.3: C1663fs | IIH |
| rs141496860 | T | C | 0.0161 | 7.13E-12 | MDN1 | midasin AAA ATPase 1 | NP_055426.1: D3304N | IIH |
| rs146069306 | A | G | 0.0200 | 8.01E-12 | FOXF1 | forkhead box F1 | NP_001442.2: G160E | PAP |
| rs529730429 | G | T | 0.0004 | 2.44E-11 | ZSWIM4 | zinc finger SWIM-type containing 4 | NP_001354763.1: L979R | IIH |
| rs200220130 | T | G | 0.0286 | 8.82E-11 | ATF6 | activating transcription factor 6 | NP_031374.2: G6E | - |
| rs546029958 | C | T | 0.0200 | 1.13E-10 | CPXM2 | inactive carboxypeptidase-like protein X2 | NP_937791.2: N171R | - |
| rs148498055 | A | G | 0.0007 | 1.18E-10 | CUZD1 | CUB and zona pellucida like domains 1 | NP_071317.2: Q497T | PAP |
| rs377424288 | A | G | 0.0084 | 1.81E-10 | RGCC | regulator of cell cycle | NP_054778.2: E26K | IIH |
| rs200588575 | G | A | 0.0175 | 2.04E-10 | CCSER2 | coiled-coil serine rich protein 2 | NP_061872.2: M334V | IIH |
| rs753766964 | C | A | 0.0258 | 9.81E-10 | NDFIP2 | nedd4 family interacting protein 2 | NP_061953.2: T102P | - |
| rs567226732 | A | G | 0.0091 | 1.05E-09 | KIF17 | kinesin family member 17 | NP_065867.2: S268L | IIH |
| rs121908162 | T | C | 0.0405 | 1.25E-09 | KIF1B | kinesin family member 1B | NP_001352881.1: T873I | IIH |
| rs140545796 | T | A | 0.0147 | 1.31E-09 | TCAIM | T cell activation inhibitor, mitochondrial | NP_776187.2: Y88C | IIH |
| rs188199560 | A | G | 0.0042 | 2.87E-09 | MSTN | myostatin | NP_005250.1: A238V | - |
| rs573651175 | A | G | 0.0063 | 4.51E-09 | SPNS2 | sphingolipid transporter 2 | NP_001118230.1: R386H | IIH |

<sup>a</sup>Contributing group (IIH or PAP) to the significance of the gene filtered by p-value <5e-07.

### Gene-based associations using SKAT and PWAS analyses

To improve the biological interpretation of the observed associations, we applied SKAT as a segment-based association method. We restricted the analysis to coding regions based on UKB genotyping imputed set (∼640k variants, see Methods). SKAT resulted in 57 genes that were reported as significant. In addition, we applied PWAS that takes into consideration the impact on the protein function of each gene for each individual (see Methods). PWAS resulted in 61 significant genes (q-value <0.05). All significant genes identified by SKAT and PWAS are listed in Supplementary **Table S5** and **Table S6**, respectively.

To increase interpretability, we focused on coherence between the methods that were based on the same input. Importantly, the different association methods (coding GWAS, PWAS, and SKAT) use the same set of variants, however the underlying models for identifying associations are substantially different. Firstly, we sought gene overlap between the genes listed by SKAT, PWAS, and coding-GWAS (**Fig. 4**). We identified FOXF1 and RGCC genes that were also among the most highly significant genes (Supplementary **Tables S5 and S6**). In addition, there were 5 genes that were shared between the SKAT and PWAS, and another 7 between the coding GWAS and SKAT. Among the shared genes of coding GWAS and SKAT are SLC28A3 which is expressed in the brain and acts in the homeostasis of endogenous nucleosides and KCNMB2 that regulates calcium activated potassium channel. Among the shared genes of PWAS and SKAT, we identified BARX2, a gene that controls the expression of neural adhesion molecules such as L1.

**Fig. 4.**
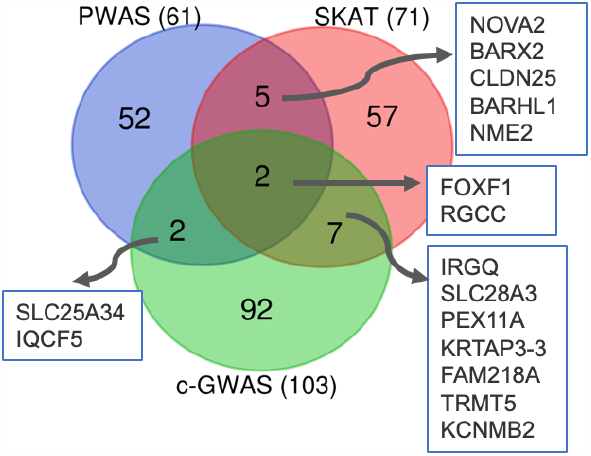
Venn diagram for the top results from SKAT (71 genes, p-value 1.0e-05), PWAS (61 genes, q-value <0.05), and coding GWAS (c-GWAS 103 genes, p-value <5e-07). The shared genes are listed. Inspecting the gene lists associated with IIH/PAP according to SKAT and PWAS showed that the lists are not particularly enriched in any specific function.

**Table 4** compiles biological knowledge for the top 10 genes from the gene-based models of SKAT and PWAS. The most significant gene (q-value: 3.0e-52) is NOVA2 (Neuro-oncological ventral antigen 2, **Table 4**). NOVA2 acts during CNS development to regulate alternative splicing. By regulating splicing, NOVA2 determines vascular morphogenesis and endothelial biology (26). Based on Open targets compilation from the GWAS catalog we observed that for many genes the genetic effects are linked to physical (e.g., BMI), neuronal and eye measurements. While only two overlapping genes by (FOXF1 and RGCC) are shared between the 10 most significant associated genes by PWAS and SKAT methods **(Table 4)**, such overlap is highly significant (p-value 1.43E-06). Interestingly, almost all genes are either known as causal for monogenic diseases (as reported by OMIM), or associated with other diseases (**Table 4**). Note that several gene-diseases concerns the retinal (NRL, RBAK, VRK3) and neurodegenerative diseases (CALML3, UBE2D3). However, some genes display pleotropic effects as indicated by the high number of gene association traits.

**Table 4.** SKAT and PWAS -the top 10 associated genes for IIH/PAP group from UKB.

| Symbol | Rank (S, P)<sup>a</sup> | Gene name | Adj. p-value (S) | # of traits<sup>b</sup> | Top GWAS trait<sup>c</sup> | Tissue enhanced (body)<sup>c</sup> | OMIM / DOID<sup>d</sup> |
| --- | --- | --- | --- | --- | --- | --- | --- |
| NOVA2 | S1 | NOVA alternative splicing regulator 2 | 3.00E-52 | M | Brain | Brain | neurodevelopment |
| FOXF1 | S2, P2 | forkhead box F1 | 5.10E-36 | H | Vasc | Intestine | capillary dysplasia |
| NME2 | S3 | NME/NM23 nucleoside diphosphate kinase 2 | 1.30E-22 | M | Phys | Brain | [cancer] |
| RGCC | S4, P1 | regulator of cell cycle | 1.60E-22 | M | Bone | Bone marrow | Kahrizi syndrome |
| NRL | S5 | neural retina leucine zipper | 2.70E-17 | M | Eye | Retina | retinal degeneration |
| DEFB104A | S6 | beta-defensin 104 | 3.30E-13 | L | Immune | Epididymis | [autoimmune diseases] |
| CALML3 | S7 | calmodulin like 3 | 1.20E-10 | M | Brain | Skin | neurodegenerative disease |
| TPRKB | S8 | EKC/KEOPS complex subunit TPRKB | 1.30E-10 | L | Brain | Low specificity | Galloway-Mowat syndrome |
| VRK3 | S9 | VRK serine/threonine kinase 3 | 5.10E-09 | - | Brain | Testis | retinitis pigmentosa |
| LSM11 | S10 | U7 small nuclear RNA associated | 4.00E-08 | H | Brain | Low specificity | Aicardi-Goutieres syndrome |
| RBAK | P3 | RB associated KRAB zinc finger | 1.95E-04 | M | Heart | Low specificity | retinal disease |
| H2AP | P4 | H2A.P Histone | 7.25E-04 | M | Eye | Testis | Galloway-Mowat syndrome |
| BCL2L2 | P5 | Bcl-2-like protein 2 | 9.15E-04 | H | Phys | Low specificity | [cancer] |
| ORC5 | P6 | origin recognition complex subunit 5 | 9.15E-04 | M | Brain | Low specificity | Meier-Gorlin syndrome |
| FUNDC2 | P7 | FUN14 domain containing 2 | 2.57E-03 | L | Blood | Tongue | Moyamoya disease |
| UBE2D3 | P8 | ubiquitin conjugating enzyme E2 D3 | 2.57E-03 | H | Blood | Low specificity | neurodegenerative disease |
| ZNF639 | P9 | zinc finger protein 639 | 2.57E-03 | L | Blood | Low specificity | [cancer] |
| MRPS6 | P10 | mitochondrial ribosomal protein S6 | 2.69E-03 | H | Mix | Choroid plexus | artery disease |

<sup>a</sup>Rank by statistics of PWAS (P) and SKAT (S). <sup>b</sup>According to OTG, the number of genetic-associated traits partitioned to low (L:1-2), medium (M:3-10), and high (H:>11). <sup>c</sup>Broad-sense phenotypes based on OTG and the highest tissue enhancement score by HPA (see Methods). Phys, physical measurements (e.g., BMI); Vasc, vascular diseases; Brain, neurological/mental condition. <sup>d</sup>OMIM and Disease ID. Groups of complex diseases are marked by parentheses.

### GWAS for IIA and PAP based on the Finnish population

The limited size of the affected group (IIH/PAP) together with the essential use of covariates for the UKB cohort (27) challenge the confidence of detection. We therefore sought the FinnGen resource that covers disjoint population of 377k. No significant results were reported from FinnGen Fz8 (Dec 2022; 197 IIH cases and 244,644 controls). However, following the increased number of individuates from a recent release (Fz9, May 2023: 231 IIH cases and 350,251 controls), a single independent variant (2:237659125:G:A; rs545417105) met the GWAS threshold (p-value <5e-08; **Fig. 5A**, red circle). This variant is very rare in all populations, with AF of 2.9e-04 in healthy Finnish population (Supplementary **Fig. S4**). Testing the relevance of rs545417105 with respect to PheWAS (28) showed that most significant phenotypes were associated with an increased risk for IIH and also PAP (**Fig. 5B**, red circles).

**Fig. 5.**
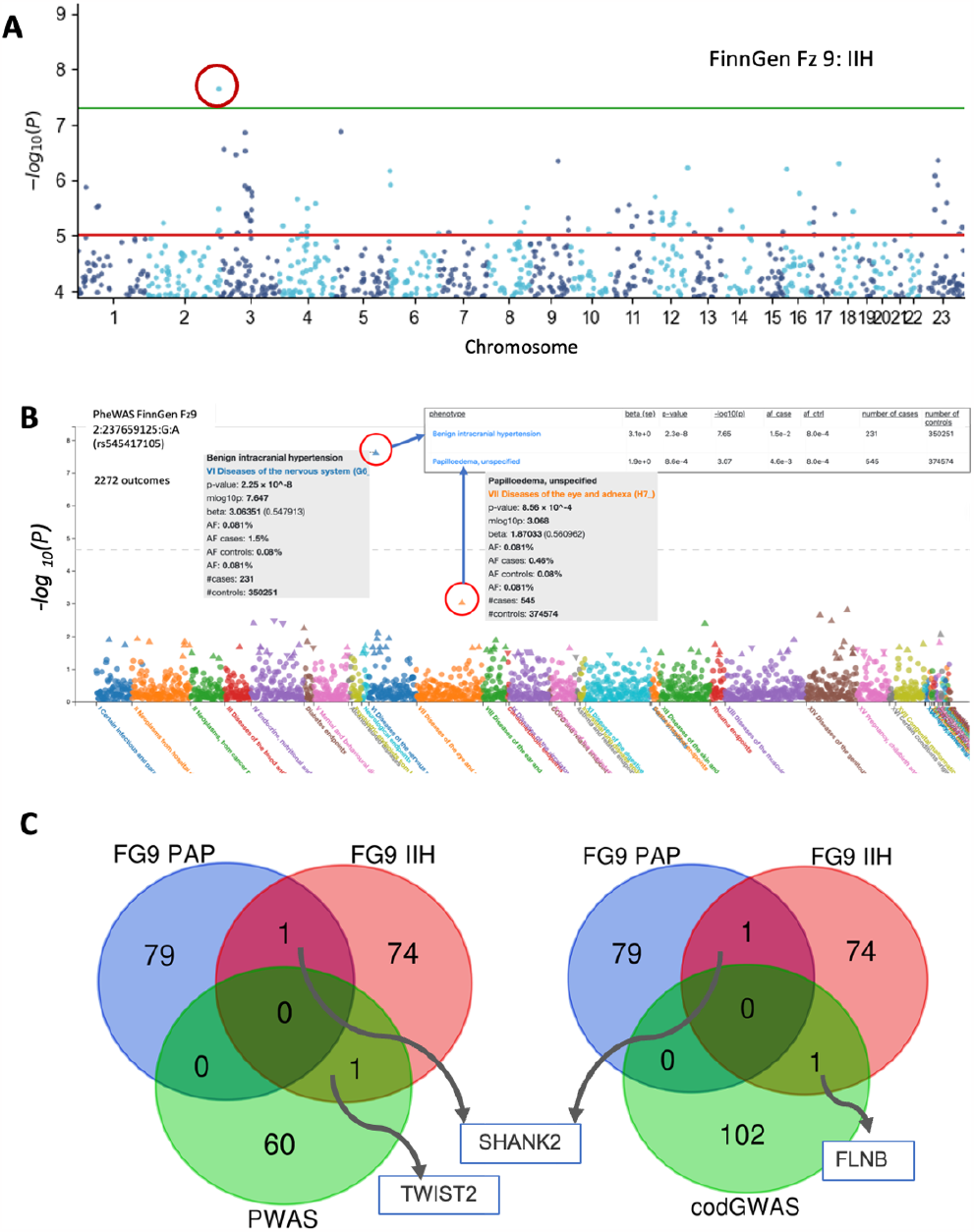
FinnGen Fz9 results for IIH associations. **(A)** Manhattan plot of the results from FinnGen Fz9 for Benign intracranial hypertension (named G6_BENINTRAHYP). The significant variant in Chr. 2 is marked by a red circle. For fine mapping analysis see Supplementary **Fig. S4. (B)** PheWAS results for variant 2:237659125:G:A (rs545417105). All phenotypes (with 2272 codes) are clustered by their medical relevance (color coded). The triangle pointing up and down indicate an increase or decrease in the risk for the relevant disease, respectively. The most significant phenotypes were associated with an increased risk for IIH and PAP. The AF ratio of the alternative allele in cases and controls is 18.75 and 5.75 for IIH and PAP, respectively. The statistical evidence is shown in the inset. **(C)** Venn diagram for FinnGen Fz9 for IIH and PAP (with p-value <1e-05) and by the inferred associated genes (based on nearest gene mapping) compared to results of PWAS (left) and coding GWAS (right) for the IIH/PAP group from UKB. For detailed comparison with more relaxed thresholds see Supplementary **Fig. S6**. FinnGen summary statistics of IIH and PAP is listed in Supplementary **Table S7**. The shared genes are indicated.

FinnGen Fz9 includes 545 cases and 374,574 controls for papilledema, unspecified (PAP). Nevertheless, none of the variants reached statistical significance of p-value <5e-08 (Supplementary **Fig. S5**). Notably, two variants are very close to the threshold: Chr8:75365414:A:G (rs144064178; p-value 6.12e-8), and Chr 4:162403766:T:C (rs1235196153; p-value 6.7e-8) (Supplementary **Fig. S5**, circles). No overlap at variant levels was observed between the FinnGen summary statistics and the results of PWAS and coding GWAS. However, the PWAS and coding GWAS overlap the FinnGen for IIH (FG Fz9, p-values <1e-05) by a single gene each, with TWIST2 and FLNB, respectively (**Fig. 5C**). Filamin B (FNLB) is detected to be shared data for IIH and coding GWAS. FLNB (filamin B) is actin-binding protein with great functional diversity and involvement in human diseases (29). In addition, SHANK2 was identified as a shared genes for FG Fz9 by applying GWAS analysis for IIH and PAP. SHANK2 plays a role in postsynaptic scaffolding, and alteration in its regulation is a known predisposition to autism or mental retardation. This association relies on a single intronic variant. Thus, an impact on this variant on the SHANK2 protein function cannot be drawn. GWAS results from FinnGen Fz9 for IIH and PAP are listed in Supplemental **Table S7**.

### Functional interpretation by external knowledge

We anticipate that all genetic association methods suffer from a shortage in detection power. This is support by the observation that the gene-based methods (SKAT and PWAS) and the coding GWAS (**Fig. 4**) share only a negligible overlap with FinnGen cohorts (**Fig. 5C**). We therefore address the limitation of large number of false positives by seeking enrichment of functional annotations. To this end, we activate the schemes of FUMA-GWAS (30) with coding GWAS (with p-value <5e-06) as input. **Fig. 6A** shows the results of the GO annotation of molecular processes (GO:MP) with enrichment of motor activity and cytoskeletal dynamic. The cellular component (GO:CC) indicates an enrichment of genes that act in ciliary fiber transition, microtubule complexes and various dynamic cytoskeletal protein complexes **(Fig. 6B)**. The enrichment of cilial genes proposes the involvement of structures that combine cytoskeletal dynamics and cell projections.

**Fig. 6.**
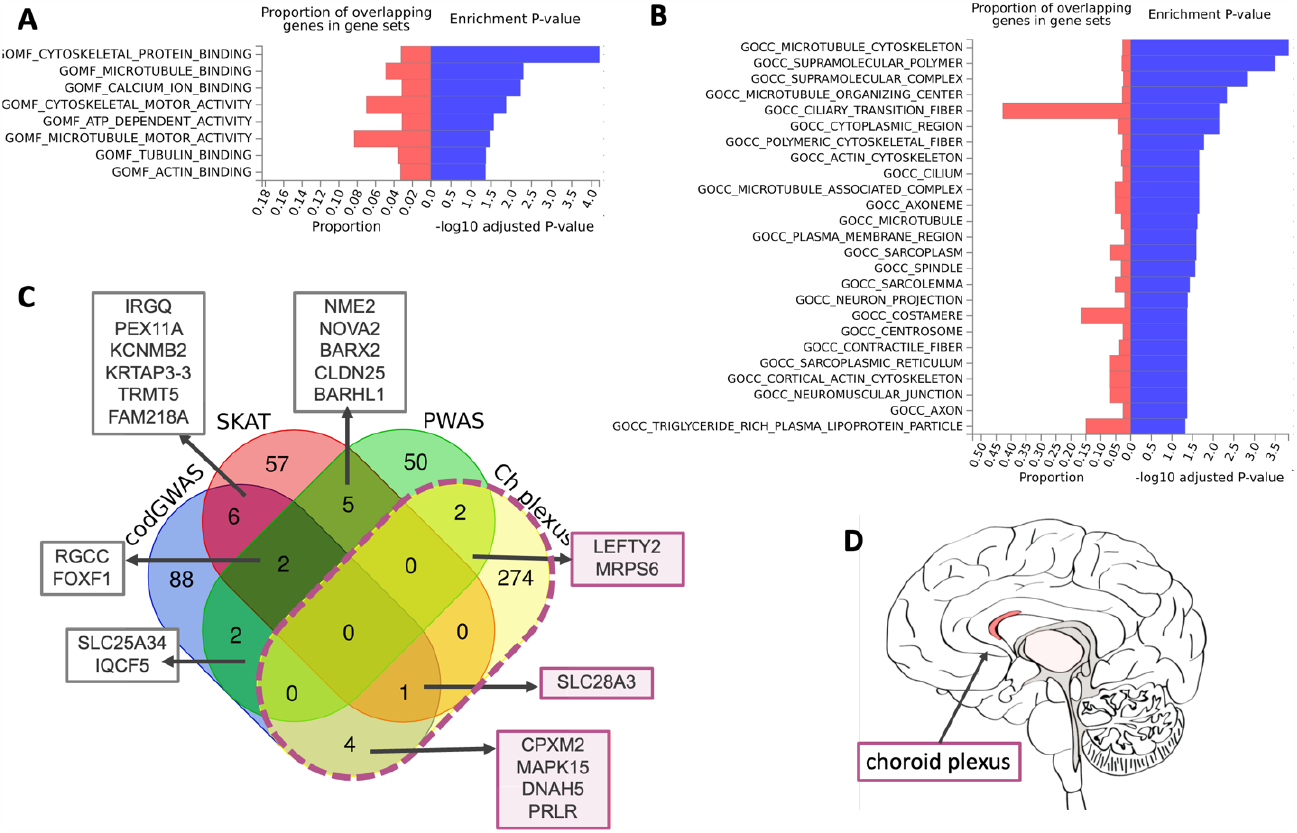
Functional enrichment of the gene lists from association studies for IIH/PAP. **(A)** Enrichment test of coding GWAS (p-value <5.0e-06) by FUMA-GWAS. Enrichment for GO (gene ontology) molecular process GOMF). **(B)** Enrichment test of by GO cellular component annotations (GOCC). In red, the proportion of overlapping genes. In blue, the enrichment statistics converted to -log10(p-value). **(C)** Venn diagram for the results from coding GWAS (103 genes, p-value <5.0e-07), PWAS with 61 genes with q-value <0.05, SKAT (71 genes, p-value <1e-05) and choroid plexus enhanced expressed genes (HPA annotated, total 281 genes, Supplementary **Table S7**). **(D)** Brain schematic expression map of gene LEFTY2 with maximal expression in choroid plexus (red) and weak expression in the thalamus, surrounded by brain ventricles (gray color).

Comparing the coding GWAS with PWAS and SKAT as gene-centric association methods identified 16 shared genes between any two of the association methods **(Fig. 6C)**. In seeking biological knowledge about the identified genes, we search for genes that act in the choroid plexus epithelium, the source of CSF and the osmotic balance in the brain ventricles. We collected a set of “tissue-enhanced” expression with the choroid plexus (HPA, human proteome atlas) (24). The overlapping genes with the enriched choroid plexus gene set (281 enhanced expression; Supplemental **Table S7**) identified seven genes (**Fig. 6C**). These genes include rs45495391 in MAPK15 (mitogen-activated protein kinase 15), rs185855852 in DNAH5 (dynein axonemal heavy chain 5), rs546029958 in CPXM2 (carboxypeptidase X, M14 family member 2) and PRLR (prolactin receptor). LEFTY2 (left-right determination factor 2) and MRPS6 (mitochondrial ribosomal protein S6) are shared with PWAS and rs141990386 in SLC28A3 (solute carrier family 28 members 3) were identified by coding GWAS and SKAT (**Fig. 6C)**. Notably, these variants are ultra-rare, and are predicted to be damaging according to variant inference tools (e.g., Polyphen2). MAPK15, DNAH5 and SLC28A3 have microtubule-related functions with an important role in cilia motility and pathology. The expression of LEFTY2 transcripts in brain tissue identified an enhanced expression specific to the choroid plexus **(Fig. 6D)**. The overlap for 7 of 16 identified gene candidates is very significant (p-value of hypergeometric test p-value 9.47e-03). Thus, the associated genes with enhances expression in choroid plexus argue for a role of cilia function as likely causal in IIH and PAP.

## Discussion

Epidemiological studies estimated that more than 90% of IIH patients are obese, and over 90% are women of childbearing age (31). Numerous factors (e.g., pregnancy, multivitamins, oral contraceptives, corticosteroids, or antibiotic use) that were proposed to be associated with IIH have been refuted by controlled studies (1). While elevated blood pressure is commonly reported in obese people, it was proposed that it is unlikely that there is a direct association between arterial hypertension and IIH. To improve patient management, effective treatment seems to require multiple experts in nutrition, neurology, endocrinology, and ophthalmology (32).

The goal of our study is to determine IIH variants/gene associations and shed light on the etiology of this complex, rare condition. The occurrence of IIH among family relatives is higher than the occurrence in the general population (33). Indirect evidence for the role of genetics in the disease’s etiology relies on the observation that IIH is much less prevalent among Asians. While it may reflect the huge difference in obesity prevalence (i.e., 33.5% in the USA compared to 3% in Japan for adult females), obesity may not be the major risk among Asians diagnosed with IIH (34). While there may be a genetic etiology accounting for the ethnic disparities, no loci or causal genes were identified (35).

In our study, we applied complementary association studies to different cohorts for identifying candidate causal genes. According to the FinnGen Fz9 cohort study for IIH, only LRRFIP1 was identified as significant. It is a cytosolic DNA sensor that has been implicated in the innate immune pathway. Altered serum or CSF levels of certain cytokines and chemokines have been demonstrated in IIH patients (36). However, we have not detected a significant immunological signature associated with IIH/PAP in the UKB cohort. Polymorphisms in LRRFIP1 positively correlate with body fat, abdominal fat, and CRP level (37). People carrying a missense rare allele in this gene (rs11680012) have 30% more abdominal adiposity relative to carriers of the major alleles (38). We show that the overlap in the the genetic signals for the studied phenotypes in FinnGen and UKB is minimal. Recall that FinnGen represents a bottleneck population (25). Moreover, association of variants to the nearest gene is prone to missed assignments. Functional studies suggested that approximately one-third of causal genes are the nearest gene to the GWAS hit (39). For our gene-centric analyses (coding GWAS, PWAS and SKAT) matching variants to genes are trivial and accurate.

Two genes showed overlap across the genetic association methodologies (coding GWAS, SKAT, and PWAS). The RGCC is expressed in almost all tissues, where it regulates cell cycle progression. It was proposed as a target for neuroprotection during the onset of Alzheimer’s disease (40). The second gene is FOXF1, a developmental gene that is among the most significant SKAT-identified genes **(Table 4)** and the top coding GWAS result. The identified variant (rs146069306, Gly160Glu) is considered deleterious and ultra-rare. FOXF1 is a hub that regulates cell adhesion, migration, and mesenchymal cell differentiation. In addition, it is a causative gene for ACDMPV (alveolar capillary dysplasia with misalignment of pulmonary veins), a disorder affecting the development of the lungs and their blood vessels (41). It is possible that, for some instances of IIH, the genetic basis is attributed to an early developmental alteration.

Under the limitation of the cohort size, we found no simple etiology for IIH/PAP. Instead, we propose a curated and ranked list of variants and genes as candidates for further investigation. We can draw some general principles from our study: (i) Almost all variants that were identified by the coding GWAS approach are rare and associated with a deleterious prediction (**Table 4**). This is in contrast to the published GWAS (study ID GCST005688) (14). (ii) IIH and PAP share a few common genetic effects (Supplementary **Tables S3, S4**). In the Finnish cohort, variant rs545417105 (LRRFIP1 gene) is strongly associated with IIH and PAP (Supplementary **Fig. 5B**). (iii) Overall agreement of different association methods, resulted in relatively low overlap among candidate genes (**Fig. 6C**). Recall that PWAS is specialized in identifying recessive inheritance, while GWAS methods focused on the statistics of additivity. (iv) The choroid plexus primary function is to produce CSF. An extended set of genes with enhanced expression levels in the choroid plexus (281 genes, HPA compilation (24)). We propose that dysregulation in CSF production or absorption can lead to IIH. There are 7 from the 16 gene candidates (**Fig. 6C**) that specify enhanced expression in the choroid plexus epithelium (p value <0.01). An attractive role of these genes in cell mobility and cilia is manifested by missense mutations in MAPK15, DNAH5 and SLC28A3. Notably, the variants associated with these genes are very rare in the general population (AF <0.02%) suggesting that these rare variants may carry an increased risk. The functionality of the cilia of the choroid plexus is an important component of brain homeostasis (42). While we anticipate many false positives in the GWAS results, the overlap with choroid plexus-enhanced genes is intriguing and propose dysregulation of this brain structure in the pathology of IIH/PAP (hypergeometric distribution p-value for the coding GWAS is 0.004).

This study has several limitations. The most important drawback is the lack of statistical power due to noisy discoveries. Moreover, while IIH occurs mostly in females, we failed to identify female-specific genetic effects due to the the limited size of the studied group. Another concern stems from the difficulty in accurate diagnosis of IIH (43). We may have misdiagnosed cases in the UKB cohort that mask the ability to extract a reliable genetic signal. Indeed, the overlap of PAP patient with IHH is only 16% in UKB while it is 48% in FinnGen, suggesting underdiagnosis.

To address the limitations, we applied relaxed statistical thresholds with caution and seek coherence by different approaches. We suggest to combine complementary association studies, to create a unified set of IIH and PAP, to analyze independent populations, and to incorporate knowledge-based methods. These has the potential to narrow down and highlight the most likely candidate genes. We claim that the overlap in findings from different association methods increases detection power, despite the use of relaxed statistical thresholds. In the rare instances of IIH/PAP, our approach will benefit personalized medicine and lead to better disease management and treatment.

## Conclusions

The comprehensive study on IIH and PAP amalgamated different genetic association methods, uncovering potentially significant genes, with high confidence for 16 genes (including FOXF1, RGCC, SLC25A34, IQCF5 and LRRFIP1). By identifying candidate genes and shedding light on their potential roles in disease mechanisms, our study lays the groundwork for potential targets and personalized management strategies. Despite inherent challenges and limitations by restricted sample size, our integrative approach illuminated shared genetic factors between IIH and PAP, particularly implicating choroid plexus dysregulation in IIH pathology.

## Data Availability

The individual-level genotype data are available under the restriction of the UK Biobank (UKB) Data Analysis Platform. The UKB application ID 26664 (Linial lab).

## List of abbreviations

AF, allele frequency; BMI, body mass index; CSF, cerebrospinal fluid; GWAS, genome-wide association studies; HPA, human proteome atlas; ICD-10, international classification of diseases, 10th revision; ICP, intracranial pressure; IIH, idiopathic intracranial hypertension; PAP, papilledema; UKB, UK Biobank.

## Data availability

All genotype-phenotype association data generated during this study are included in this published article and its Supplementary information Tables. The individual-level genotype data are available under the restriction of the UK Biobank (UKB) Data Analysis Platform. The UKB application ID 26664 (Linial lab).

No custom code was developed for this project. We used in-house developed pipelines for the analytic procedures. The computational pipeline for processing the UKB data is open source and available at https://github.com/nadavbra/ukbb_parser (through both Python and a command-line interface).

## Acknowledgments

We thank the reviewers for useful comments and suggestions. We thank the Linial lab for fruitful discussions.

## Supplementary Figures

**Fig. S1**. Distribution of BMI by sex.

**Fig. S2**. Venn diagrams for PAP, IIH and unified group (both) for all GWAS hits (genes and variants)

**Fig. S3**. Venn diagram for coding GWAS results for IIH, PAP and the unified group (both)

**Fig. S4**. Manhattan plot according to FinnGen Fz9 for IIH and fine mapping

**Fig. S5**. Manhattan plot according to FinnGen Fz9 for PAP and fine mapping

**Fig. S6**. Venn diagram of PAP, IIH from FinnGen Fz9 and coding GWAS hits (genes and variants)

## Supplementary Tables

**Table S1**. U-test statistics for demographic data; support Table 1.

**Table S2**. List of comorbidities from UKB and FinnGen populations; source for Table 2.

**Table S3**. List of significant variants from all GWAS for IIH/PAP; source for Fig. 2.

**Table S4**. List of significant variants from coding-GWAS; source for Fig. 3, Table 3.

**Table S5**. List of significant genes from SKAT; source Figs. 4-6, Table 4.

**Table S6**. List of significant genes from PWAS; source for Figs. 4-6, Table 4.

**Table S7**. List of enhanced expression in choroid plexus; source for Fig. 6.

## Ethical approval

The study was approved by the University Committee for the Use of Human Subjects in Research Approval number 12072022 (July 2023).

## Author Contributions

ML, EB and IM designed the study. IM and ZD contributed the clinical perspective. RZ and MK contributed to data extraction, statistical analyses and interpretation. ML wrote the initial version. All authors read and commented for the final manuscript.

