## Supplemental Fig. S for "The genetics of idiopathic intracranial hypertension (IIH): Integration of population studies and clinical data"

### Supplemental Figures

**Fig. S1.** Distribution of BMI by sex.
**Fig. S2.** Venn diagrams for PAP, IIH and unified group (both) for all GWAS hits (genes and variants)
**Fig. S3.** Venn diagram for coding GWAS results for IIH, PAP and the unified group (both)
**Fig. S4.** Manhattan plot according to FinnGen Fz9 for IIH and fine mapping
**Fig. S5.** Manhattan plot according to FinnGen Fz9 for PAP and fine mapping
**Fig. S6**. Venn diagram of PAP, IIH from FinnGen Fz9 and coding GWAS hits (genes and variants)

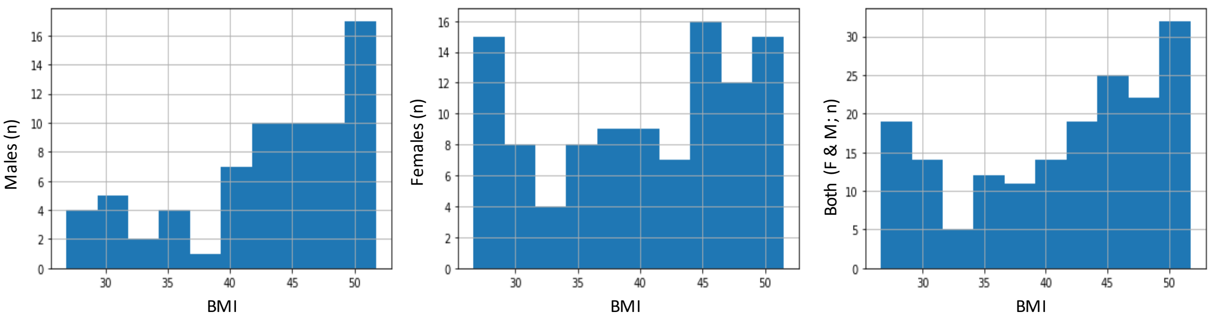


**Fig. S1.** BMI distribution of males, females and both. The mean and standard deviation are shown in Table 1.

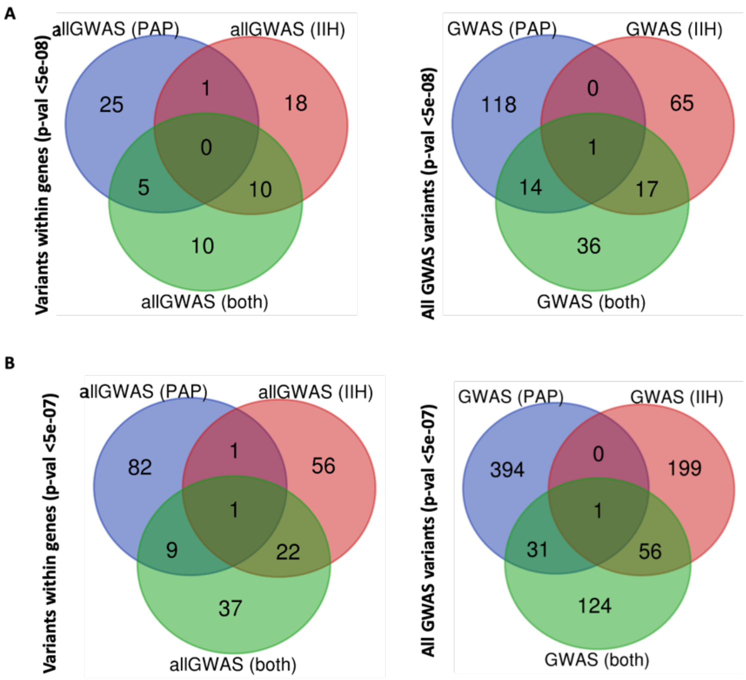


**Fig. S2.** Venn diagram for all GWAS results for IIH, PAP and the unified group (both; IIH/PAP). **(A)** GWAS threshold of p-value <5e-08 by genes based on variants within genes (left) and all variants (right). Note that VLDLR gene is shared by all three groups. **(B)** GWAS threshold of p-value <5e-07 by genes based on variants within genes (left) and all variants (right). The variants and the genes and their identity according to the Venn diagrams are listed in Supplementary **Table S3**.

NXPH1 is shared between PAP and IIH group (however, the association is based on different variants for each group). Specifically, only a single variant for the IIH group and 61 variants of NXPH1 in PAP group. Only a single intergenic variant (rs12203577) is shared by all three groups. This SNP is associated with p-values of e-37 to e-70, with negative z-values. The results of the overlapping groups are compiled in supplemental **Table S3.**

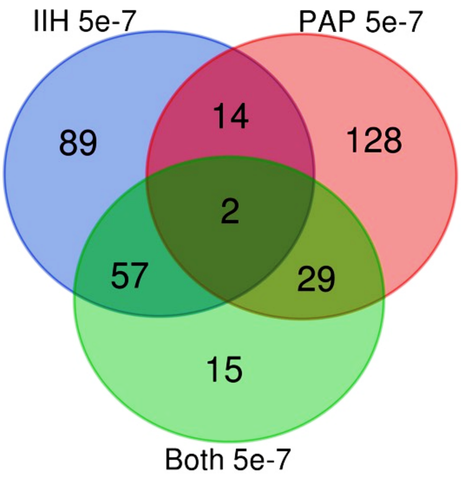


**Fig. S3.** Venn diagram for coding GWAS results for IIH, PAP and the unified group (both; IIH/PAP). Coding GWAS threshold was set to <5e-05. There are 16 shared genes but only 2 are recovered by using the combined cohort (IIH/PAP). These genes are IMPACT and PCDHGA1.

The other 14 genes that are shared between IIH and PAP are: PAM, UBN1, SBSPON, SUSD6, CFAP206, RREB1, TOX, KIF5B, CLEC4F, VPS53, CACNA1S, PLK5, HSF1, and COLQ. The variants and the genes and their identity according to the Venn diagrams are listed in Supplementary **Table S4**.

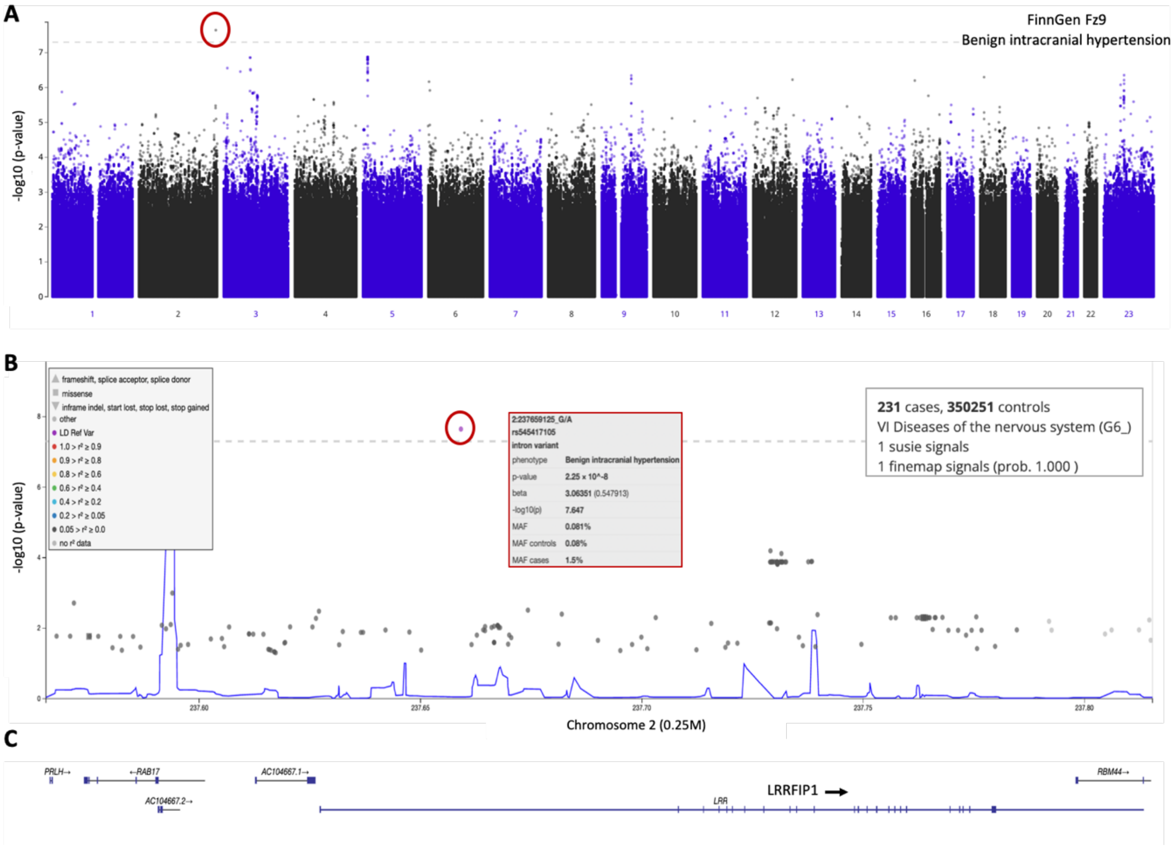


**Fig. S4.** FinnGen results for IIH associations. **(A)** Manhattan plot of the results from FinnGen Fz9 for all reported variants (21 M) for Benign intracranial hypertension (named G6_BENINTRAHYP). The variant 2:237659125:G:A (rs545417105) reached significance with a p-value of 2.25e-08 (circled red). The analysis was performed on 231 cases and 350,251 controls**.** The dashed horizontal line marks the p-value threshold of 5.0e-08 for GWAS. **(B)** Fine map of variant 2:237659125:G:A (rs545417105) for a segment of 250k in Chr 2. The candidate SNP is associated with the statistical measures (red circles). Each imputed SNP is marked if the pair correlation $r^2$ is <0.05. The blue plot is an estimate for the recombination rate from the LD blocks. **(C)** Chromosomal segment of Chr2 from position 237.57 M to 237.82 M (total 250k). Gene lengths are shown by a horizontal line with the exons marked by the vertical lines. The arrow denotes LRRFIP1 transcription directionality.

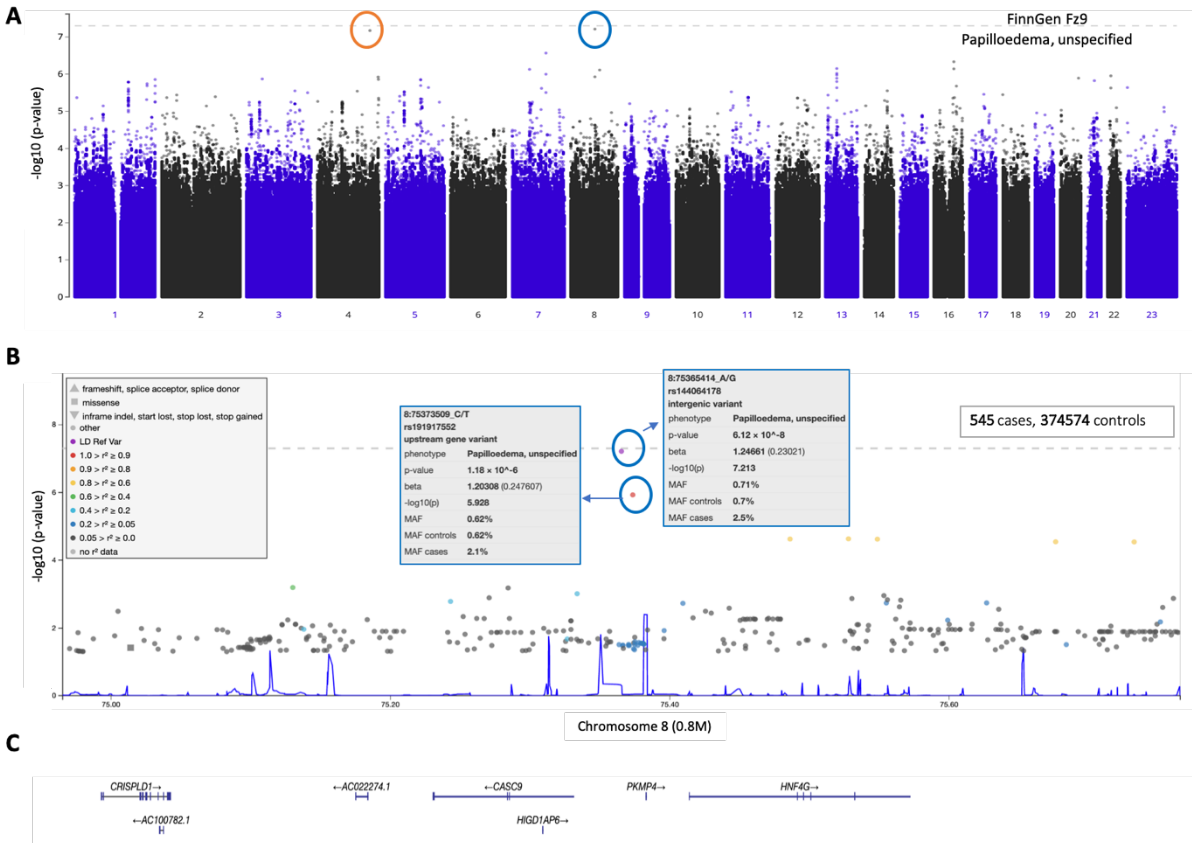


**Fig. S5.** FinnGen results for PAP associations. **(A)** Manhattan plot of the results from FinnGen Fz9 for papilloedema, unspecified. The variants 4:162403766:T:C (rs1235196153) and 8:75365414:A:G (rs144064178) are circled in orange and blue, respectively. Both variants are slightly below the significance threshold dashed horizontal line). **(B)** The fine mapping analysis for a segment of 1.6M in Chr 8. The candidate SNPs are associated with the statistical measures (blue circles). All imputed SNP are shown if the pair correlation ($r^2$) is <0.05. The blue plot is an estimate for the recombination rate (see Methods). **(C)** Chromosomal segment of Chr8 from position 75.0 M to 75.8 M (total 0.8M). Gene lengths are shown by horizontal lines with exons marked by the vertical lines. The arrows indicate transcription directionality. The nearest gene of the leading variant in Chr8 is HNF4G.

A

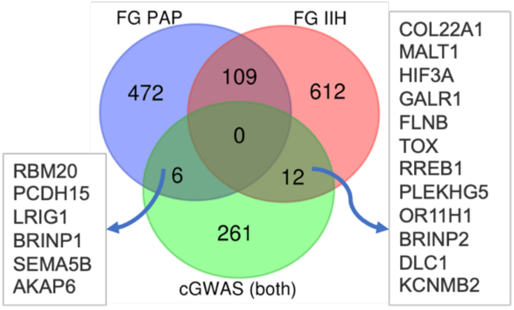


| Groups | # of genes | # of unique genes |
|---|---|---|
| FG IIH | 777 | 733 |
| FG PAP | 618 | 587 |
| cGWAS (both) | 293 | 279 |

B

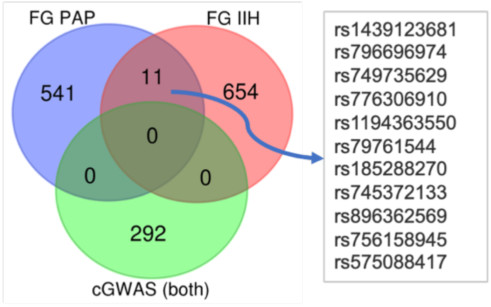


| Groups | # of SNP ID | # of unique SNP ID |
|---|---|---|
| FG IIH | 689 | 665 |
| FG PAP | 566 | 552 |
| cGWAS (both) | 293 | 292 |

**Fig. S6.** Venn diagram for FinnGen Fz9 summary statistics for IIH and PAP and coding GWAS for IIH/PAP group. **(A)** GWAS results for IIH and PAP by the inferred associated genes (based on nearest gene mapping). The overlap of IIH and PAP is significant (hypergeometric test p-value 3.2e-42). The overlap of the gene list with coding GWAS is minimal. The number of elements used for the analyses are specified (Bottom). **(B)** GWAS results for IIH and PAP by variants IDs. There is no overlap with coding GWAS and only minimal overlap between the IIH and PAP associated variants. The number of elements used for the analyses are specified (Bottom). The FinnGen summary statistics of IIH and PAP is listed in Supplementary **Table S7**.
